# Impact of Antibody Cocktail Therapy Combined with Casirivimab and Imdevimab on Clinical Outcome for Covid-19 patients in A Real-Life Setting: A Single Institute Analysis

**DOI:** 10.1101/2021.10.10.21264589

**Authors:** Yasutaka Kakinoki, Kazuki Yamada, Yoko Tanino, Keiko Suzuki, Takaya Ichikawa, Naoki Suzuki, Go Asari, Ai Nakamura, Shin Kukita, Akito Uehara, Seisuke Saito, Shohei Kuroda, Hidemitsu Sakagami, Yuuki Nagashima, Kae Takahashi, Satoshi Suzuki

**Author notes:** **Corresponding author:** Yasutaka Kakinoki, MD, PhD., Asahikawa City Hospital, 1-1-65, Kinseicho, Asahikawa, 070-8610, Japan. **Shared co-first authorship:** Dr. Yasutaka Kakinoki and Dr. Kazuki Yamada contributed equally to this article.

## Abstract

**Background:** Recent data from clinical trial suggest that antibody cocktail therapy, a combination of the monoclonal antibodies casirivimab and imdevimab, has been shown to rapidly reduce the viral load and markedly decrease the risk of hospitalization or death among high-risk patients with coronavirus disease 2019 (Covid-19). However, it remains unclear how effective in a real-life clinical setting the therapy is.

**Methods:** We retrospectively analyzed mild to moderate Covid-19 patients with one or more high-risk factors for severe disease who consecutively underwent the antibody cocktail therapy of the disease in our institute in June 2021 through early September 2021, compared to those with high-risk factors who were isolated in non-medical facilities consecutively during the same period, thereby being not given the antibody cocktail therapy there. The key outcome was the percentage of patients with Covid-19-related deterioration which needed additional medical interventions, such as oxygen support or other antiviral therapies.

**Results:** Data from 55 patients with initially receiving antibody cocktail therapy and 53 patients with isolation into non-medical facilities are analyzed. 22 (41.5 %) of 53 patients with isolation facilities were finally hospitalized to receive medical interventions. On the other hand, 13 (23.6 %) of 55 patients with antibody cocktail therapy in our hospital subsequently underwent further medical interventions because of the progression. In multivariate analysis with variables of age, BMI, and high-risk factors, the antibody cocktail therapy significantly reduced 70 % in the need for further medical interventions compared to the initial isolation in the non-medical facilities (odds ratio=0.30, 95%CI [0.10-0.87], p=0.027). Furthermore, patients with 96% or above of SPO2 were significantly more favorable for the therapy than those with 95% or below of SPO2.

**Conclusion:** The treatment of antibody cocktail was closely linked to reduction in the need for further medical interventions. The result indicates that the antibody cocktail therapy is associated with reducing the strain on hospitals, which is related to the improvement of medical management for public health care in Covid-19 pandemic era.

## INTRODUCTION

Coronavirus disease 2019 (Covid-19), caused by severe acute respiratory syndrome coronavirus 2 (SARS-CoV-2), was first detected in December 2019 and was declared a global pandemic in March 2020 [1-3]. In Japan, as of September 22, approximately 1.68 million people have been infected and 17 thousand are dead, and about 55% people of the population are fully vaccinated [4]. Although those records regarding infected figures appear to be less than in Western countries, the several issues in hospitals are challenging because of less enough hospital beds availability and shortages of hospital staffs, mostly nurses. To resolve these issues, Japan’s Ministry of Health, Labour and Welfare (MHLW) has approved the antibody cocktail of casirivimab and imdevimab, brand name of Ronapreve™ (provided from Roche globally, and from CHUGAI Pharmaceutical in Japan), for the treatment of mild to moderate Covid-19 patients with high-risk factors via intravenous infusion, which was granted a Special Approval Pathway under article 14-3 of the Pharmaceuticals and Medical Devices Act on July 19 in 2021 [5]. The approval of the MHLW has based on results from the global phase 3 REGN-COV2067 trial [6] in high-risk non-hospitalized Covid-19 patients, which showed that the cocktail therapy reduced hospitalization or any-caused death by 70% and Covid-19-related symptom duration by 4 days as well as a phase 1 clinical study with safety, tolerability and pharmacokinetics in Japanese people [7]. In addition, the ability of the cocktail to retain activity against emerging variants including delta variant has been demonstrated in vitro study [8-10]. However, these results have been coming up from randomized clinical trials and experimental studies, and therefore what is happening in a daily clinical practice on the cocktail therapy remains to be seen. Here, we describe clinical benefits of Ronapreve coming from a real-life clinical practice in our institute.

## METHODS

### PATIENTS

Eligible patients were 20 years of age or older, with a confirmed SARS-CoV-2 infection by real-time reverse transcription PCR (RT-PCR) test, a Covid-19-related fever (≧ 37.5°C) in mild to moderate condition, and presence of risk factors meeting the criteria for severe Covid-19 [6] during June 2021 through early September 2021. Local public health center made allocation decisions based on factors such as severity and urgency to deliver patients to hospitals or non-medical facilities for isolation. The use of patient’s clinical information was approved by the Research Ethics Committee of Asahikawa City Hospital which oversaw the study conduct and documentation, and the data available from non-medical facilities were authorized to be provided with fully anonymous condition by the chief officer of the local public health center. This study was conducted in accordance with the principles of the Declaration of Helsinki.

### MEDICAL INTERVENTION

Patients assigned to our institute were firstly reviewed to be applicable for the use of ronapreve according to the criteria [6]. Ronapreve was given at equal doses of 600mg of casirivimab and imdevimab combined in a 100ml normal saline solution through intravenous infusion over 30 minutes, if applicable (ronapreve group). Afterward, if necessary, patients can receive additional further therapy such as oxygen support, steroids, or antiviral drugs. Patients assigned to non-medical facilities were under watch-and-wait situation to see if they progress to the point they need hospitalization (watchful observation group). When patients possibly progress or got progressed, they were immediately transferred to hospitals to receive some treatments for Covid-19.

### KEY and OTHER OUTCOMES

Key outcomes were designated to the difference between ronapreve and watchful observation groups in terms of the necessity of additional further treatment such as oxygen support or antiviral drugs. In ronapreve group, the addition of further treatment after ronapreve administration indicated the failure of the cocktail therapy as a definition. In watchful observation group, the transfer of patients to hospitals to receive some therapies indicated that patients were under intractable or deteriorating conditions. Other outcomes were designated in ronapreve group to investigate the duration of fever and adverse events after ronapreve administration.

### STATISTICAL ANALYSIS

Logistic regressioin models for multivariate analysis were applied to evaluate the proposed significant factors in terms of the efficacy of ronapreve, using age, BMI, high-risk factors, and Percutaneous oxygen saturation (SPO2) as explanatory variables. Receiver Operating Characteristic (ROC) curves were used to determine the cut-off value for SPO2. All p-values were two sided and P-values of 0.05 or less were considered statistically significant. All statistical analyses were performed with EZR version1.50 (Saitama Medical Center, Jichi Medical University, Saitama, Japan), which is a graphical user interface for R (The R Foundation for Statistical Computing, Vienna, Austria). More precisely, it is a modified version of R commander designed to add statistical functions frequently used in biostatistics [11].

## RESULTS

### PATIENT’S CHARACTERISTICS

Patients were collected consecutively during the period from June 2021 through early September 2021, when the delta variants were widely spreading in our community. As a result, 55 patients were given Ronapreve first in our institute (ronapreve group) and 53 patients were initially assigned to non-medical isolation facilities to see the situations watchfully (watchful observation group) (Table 1).

**Table 1.**
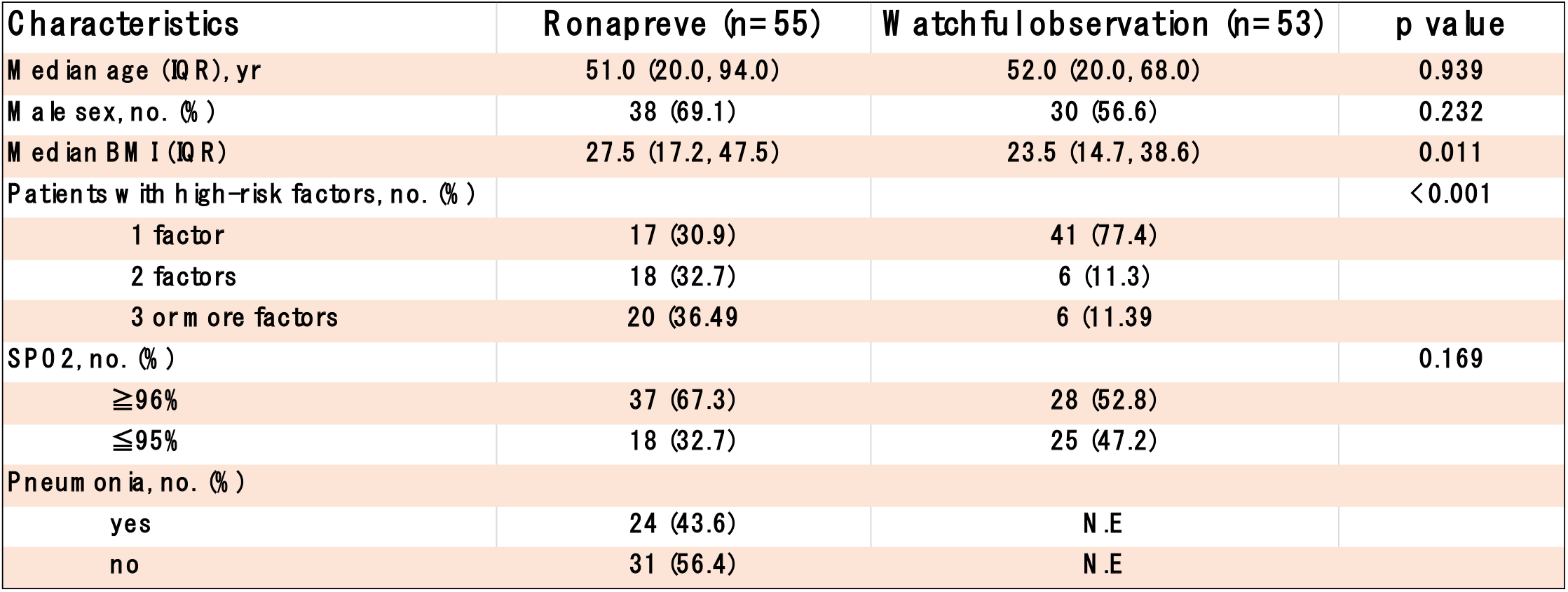
Patients’ Demographic and Epidemiological Characteristics. Abbreviations: IQR, interquartile range; BMI, body mass index; SPO2, Percutaneous oxygen saturation. #High-risk factors for severe Covid-19 include an age of more than 50 years, obesity (BMI ≧30), cardiovascular disease (including hypertension), chronic lung disease (including asthma), chronic metabolic disease (including diabetes), chronic kidney disease (including receipt of dialysis), chronic liver disease, and immunocompromise.

In ronapreve group, the median age was 51 years, 69.1% were male, and median BMI score was 27.5. In proportion of the number patients have in high-risk factors, 30.9% possessed 1 factor, 32.7% 2 factors, and 36.4% 3 or more factors. SPO2 tests revealed 67.3% of patients to be 96% or above and 43.6% of patients have pneumonia detected by CT scan, X-ray imaging, or stethoscopic findings. In contrast, in watchful observation group, although distributions of age and gender were similar, proportion of high-risk factors and BMI scores regarding disease progression were significantly lower (p<0.001 and p=0.01, respectively), compared to ronapreve group. Details of the high-risk factors patients have in both groups together are provided in the Supporting Information (Table S1).

### CLINICAL EFFICACY

#### Key outcomes

In ronapreve group, 23.6% (13/55) of patients eventually needed further medical interventions after ronapreve administration, such as oxygen support or antiviral drugs (Figure 1). However, no deterioration was found beyond 5 days after ronapreve administration, meaning the remaining 76.4% (42/55) of patients in this group fully recovered from Covid-19 (Figure 1A). On the other hand, Patients in watchful observation group showed that those who got progressive were increasingly transferred to hospitals until 12 days after the disease onset, finally mounting up to 41.5% (22/53) of patients (Figure 1B). In multivariate analysis with age, BMI, and high-risk factors as explanatory variables, ronapreve significantly reduced the need for additional treatments by 70% compared to watchful observation group patients (odds ratio=0.301, 95%CI [0.104-0.869], p=0.026) (Figure 2). Furthermore, in ronapreve group, patients with 96% or above of SPO2, the cutoff value was established by ROC curves, showed 97% reduction significantly on the additional treatment compared to patients with 95% or below of SPO2 (odds ratio=0.03, 95%CI [0.01-0.22], p<0.001) (Figure 3).

**Figure 1.**
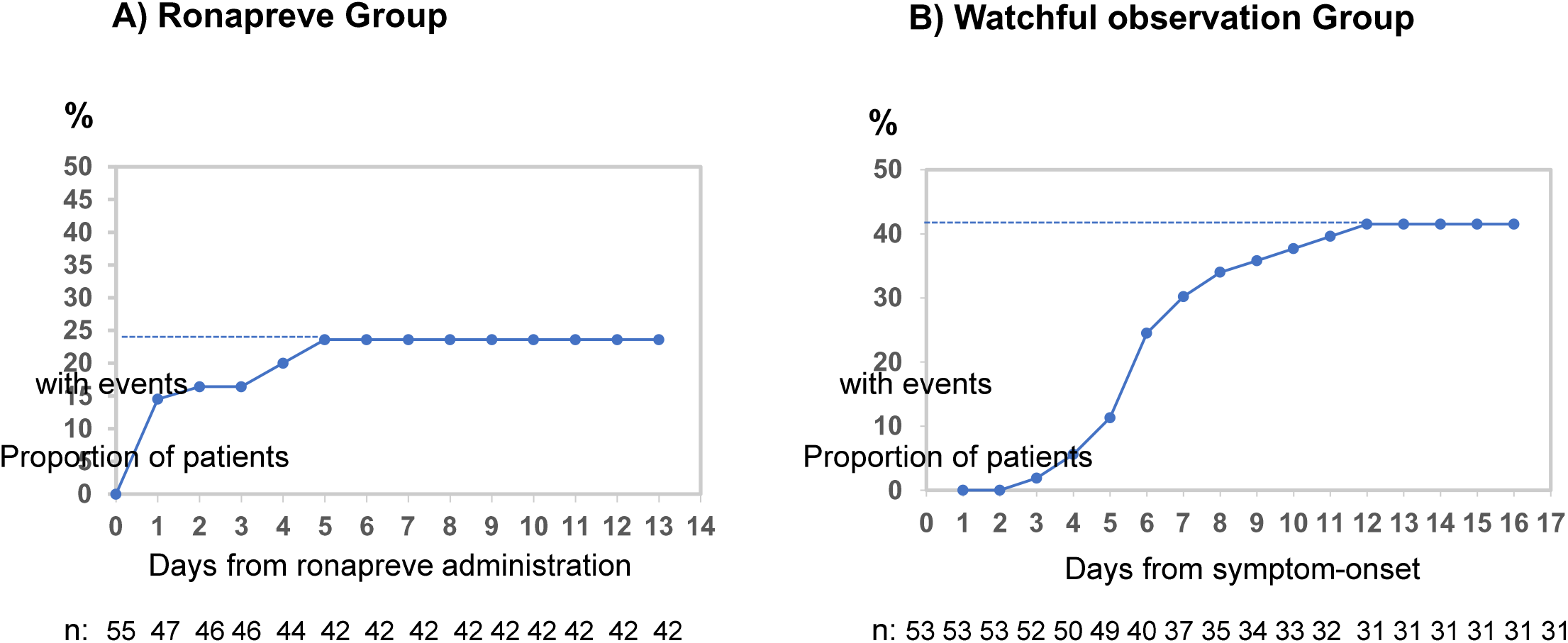
Time to additional treatment. Event indicates that additional treatments are started. A) shows time (day) to the next additional treatment after ronapreve administration (date of the dripping indicates day 0). B) shows time (day) to be hospitalized to receive medical interventions from disease onset.

**Figure 2.**
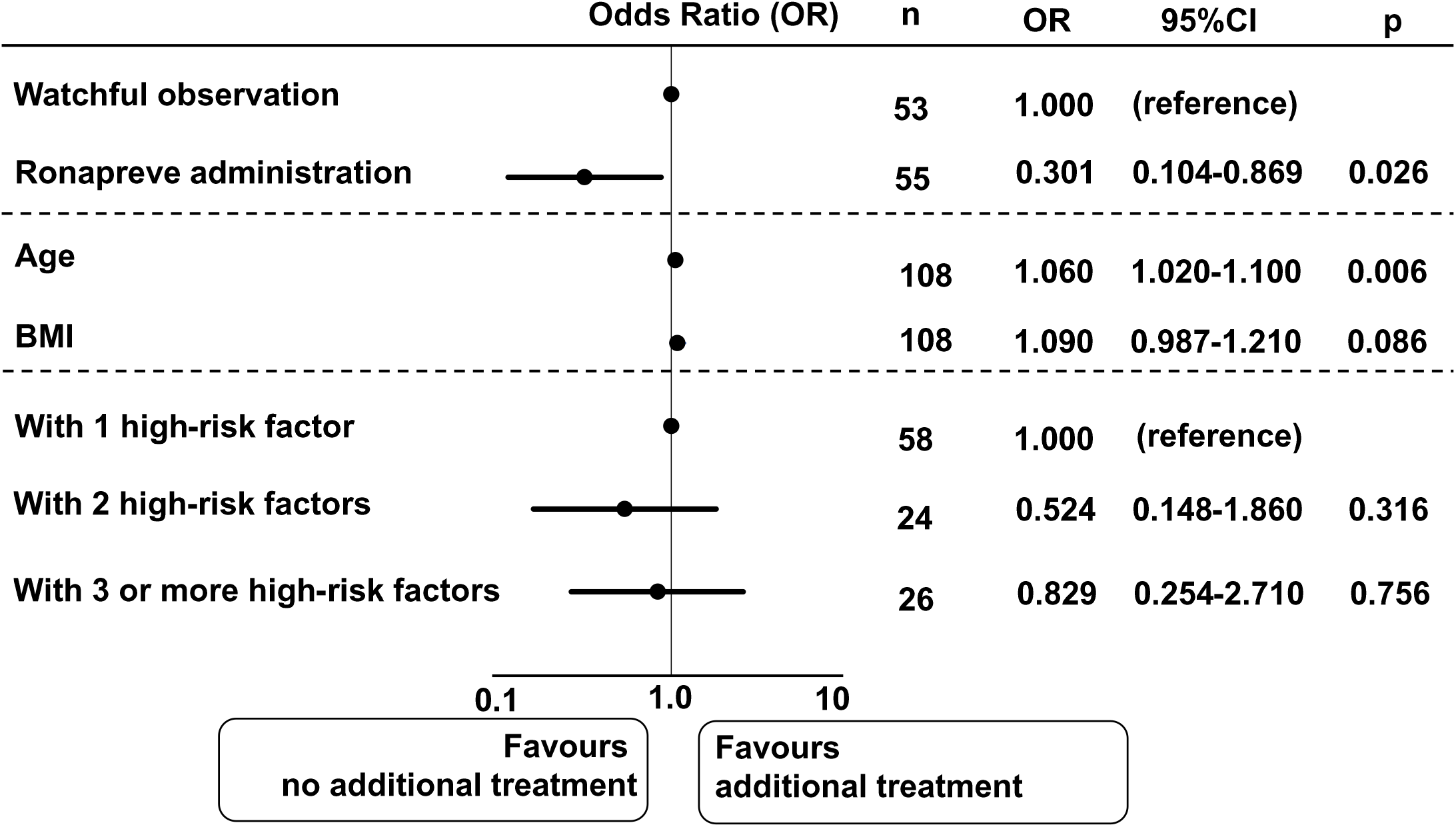
Multivariate analysis for the efficacy of ronapreve using variables of age, BMI, and high-risk factors. Forest plots depict the comparison of the incidences between watchful observation and ronapreve groups.

**Figure 3.**
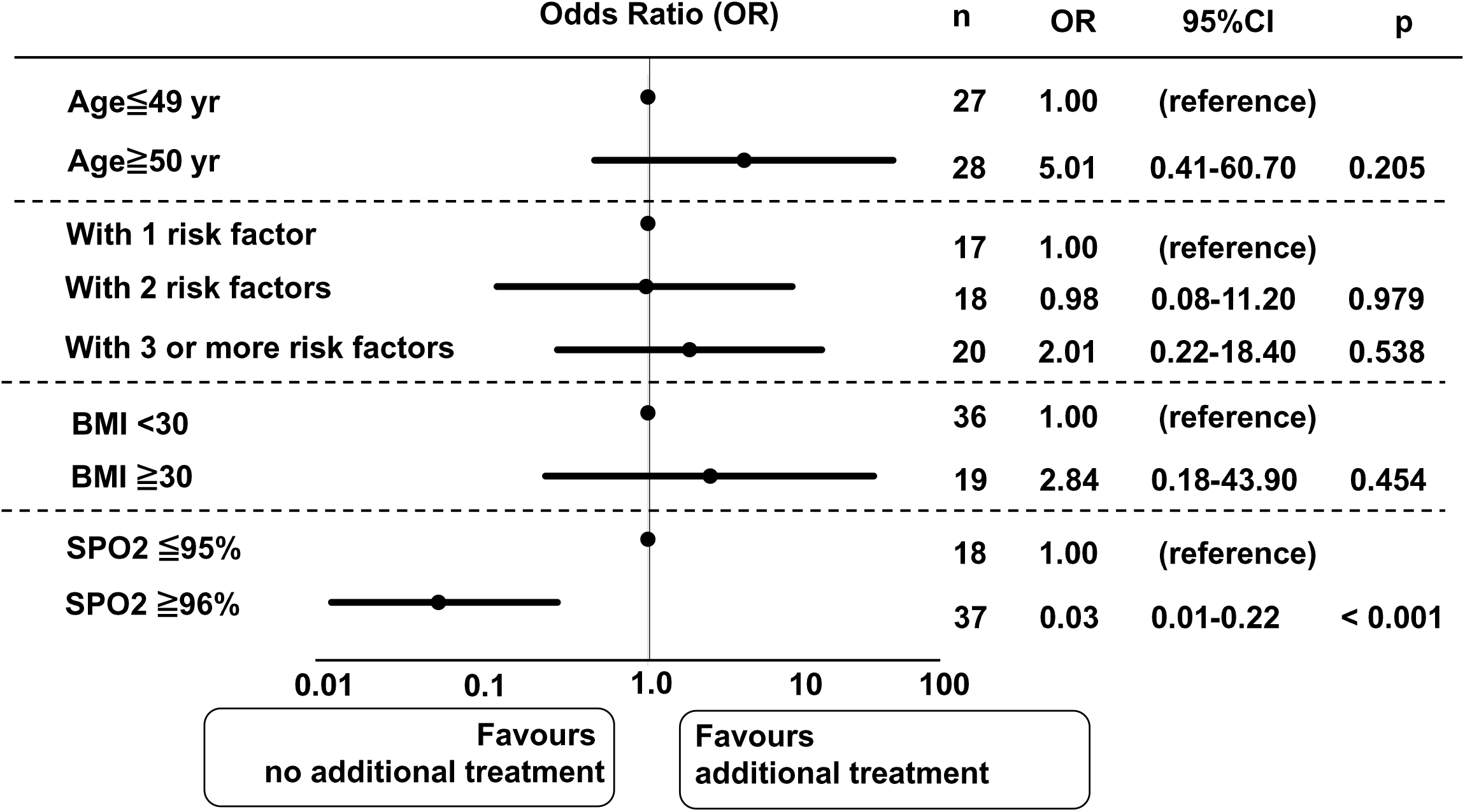
Multivariate analysis for age≧50 years, 1, or 2, or 3 or more high-risk factors, BMI≧30, and SPO2≧96% in ronapreve group. Forest plots depict the comparison of the incidences.

#### Other outcomes

Ronapreve was started at a median time of 3 days from the onset (range; 0 – 7 days) (Figure S1). Ronapreve treatment was associated with quick relief from Covid-19-related fever. Out of 27 in-patients, 14 patients (51.9%) were reduced from fever until the next day, and all 27 patients has achieved afebrile state until 4 days from the administration, though this result came from limited numbers of 27 in-patients because of missing data from other 28 out-patients (Figure 4). In regard to vaccinated patients enrolled, in 3 with one shot and in 5 with 2 shots, one and 4 patients showed to be related to the fever-down, respectively (Table S2). In aspect of adverse events, one patient showed infusion reaction of mild swelling of eyelids and urticaria of upper arms during the drip of ronapreve, resulting in stopping on the way, and 2 patients showed skin eruption around 2-3 hours after the administration (Table S3).

**Figure 4.**
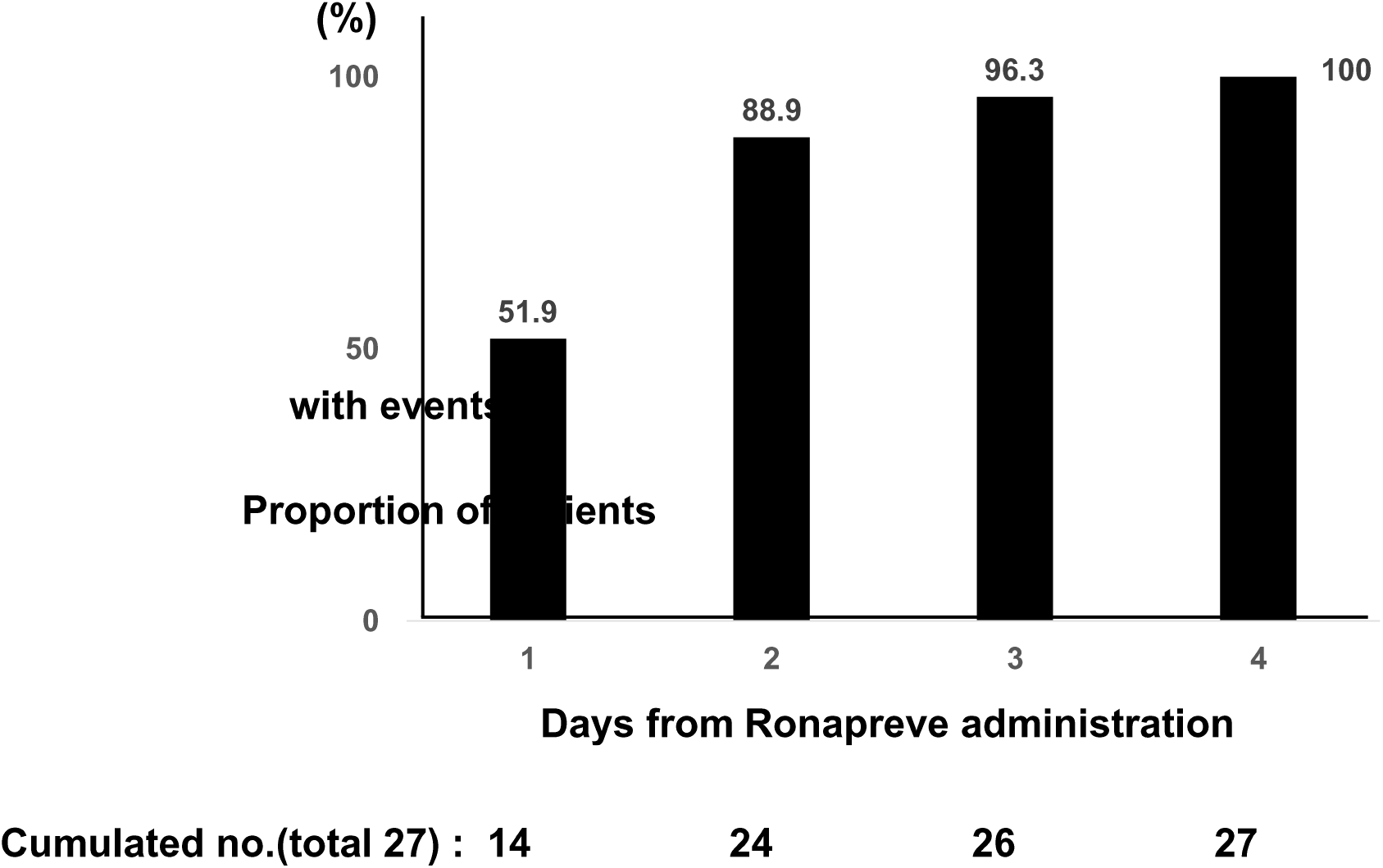
Accumulation of patinets with events from ronapreve administration (date of the dripping indicates day 0). Event indicates that fever is down.

## DISCUSSION

Ronapreve (also known as REGEN-COV in clinical trials) is a cocktail made up of two noncompeting neutralizing human IgG1 monoclonal antibodies, casirivimab and imdevimab, that target the receptor-binding domain of the SARS-CoV-2 spike protein, thereby preventing viral entry into human cells through the angiotensin-converting enzyme 2 (ACE2) receptor [8,12]. This cocktail therapy retains neutralization potency against circulating SARS-CoV-2 variants of concern, including B.1.1.7 (or alpha), B.1.351 (or beta), B.1.617.2 (or delta) and so forth in vitro and in vivo [8-10].

In a real-life practice setting, we described that the administration of ronapreve predicted 70% reduction in turning into additional treatment, as compared with watchful observation in isolation facilities where patients are under watch-and-wait situation to see if they progress to the point they need hospitalization to receive treatments (Figure 2). Moreover, we showed 97% reduction of additional treatments in those patients who were given under conditions of 96% or above of SPO2, compared to those with 95% or below (Figure 3). This result may be associated with suppression of SARS-CoV-2 itself by ronapreve before a surge of inflammation in lungs. In addition, ronapreve was related to substantially speed up recovery from Covid-19-related fever at a median time of just 1 day from the administration (Figure 4), which probably represents an additional benefit for Covid-19 patients. It is worth noting that patients in ronapreve group were under the worse conditions than those in watchful observation group in terms of risk factors for the disease progression.

Our data in a real-life setting suggest that ronapreve has the potential to prevent mild to moderate Covid-19 patients with high-risk factors from receiving additional treatments, such as supplemental oxygen, dexamethasone, or antiviral therapies because of disease progression, which should be exclusively manipulated in hospital in Japan. This result indicates that ronapreve is associated with reducing the burden of the need to take care of Covid-19 patients in hospital beds, which is related to retaining public health care resources to be normal.

Overall, our findings described above are well consistent with data from previous clinical trials regarding ronapreve. The phase 1/2 trial data showed that REGEN-COV for Covid-19 patients lowered viral load, reduced the need for medical attention, and was highly suggestive of a reduced risk for hospitalization [13]. The phase 3 clinical trial confirmed that early treatment with REGEN-COV in outpatients with high-risk factors for severe Covid-19 dramatically reduced the risk of hospitalization or all-cause death by 70.4% and the symptom duration by 4 days at equal doses of 600mg of casirivimab and imdevimab [6]. In addition, the phase 3 trial for the prevention of Covid-19 in household contacts of infected individuals showed that subcutaneous administration of REGEN-COV reduced the risk of symptomatic Covid-19 infections by 81.4% [14]. These findings suggest that REGEN-COV therapy in outpatients with Covid-19 has the potential to improve patient outcomes and substantially reduce the health care burden by lowering morbidity and mortality.

The important point for ronapreve treatment we mentioned above is that the efficacy for patients is diminished under the condition of 95% or below of SPO2. Since low SPO2 or pneumonia is probably associated with cytokine-storm-induced hyper-inflammation caused by SARS-CoV-2, steroid such as dexamethasone will be more effective to suppress the storm rather than ronapreve, because the antibody treatment probably affects the virus itself.

In conclusion, ronapreve, also known as REGEN-COV, is thought to be closely linked to reduction in the risk of hospitalization or the need for additional treatment, along with a potential benefit of prompt recovery from Covid-19-related fever. Although our data provided from a daily practice is small-sized and limited, the antibody cocktail therapy in early phase of the disease suggests a promising way to minimize the serious impact of Covid-19 on the public health care system.

## Data Availability

All data produced in the present study are available upon reasonable request to the authors.

## ACKNOWLEGEMENT

The authors thank all the participating physicians and co-medical staffs for their care of patients with Covid-19.

## SUPPORTING INFORMATION

Additional supporting information may be found online in the Supporting Information section at the end of the article.

## REFERENCES

1. Zhu N, Zhang D, Wang W, et al. A novel coronavirus from patients with pneumonia in China, 2019. N Engl J Med 2020; 382: 727–733.

2. World Health Organization. WHO Director-General’s opening remarks at the media briefing on COVID-19 – 11 March 2020. 2020 (https://www.who.int/director-general/speeches/detail/who-director-general-s-opening-remarks-at-the-mrdia-briefing-on-covid-19---11-march-2020).

3. Wu F, Zhao S, Yu B, et al. A new coronavirus associated with human respiratory disease in China. Nature 2020; 579: 265–269.

4. Ministry of Health, Labour and Welfare. Current status of the novel coronavirus infection and the response of the MHLW. 2020. Available at: https://www.mhlw.ga.jp/stf/newpage_12312.html. Accessed 22 September 2021.

5. Ministry of Health, Labour and Welfare. Approval for RonapreveTM (casirivimab and imdevimab) for the treatment of patients with mild to moderate Covid-19. Available at: https://www.mhlw.ga.jp/stf/newpage_19940.html. Accessed 20 July 2021.

6. Weinreich DM, Sivapalasingam S, Norton T, et al. REGEN-COV antibody cocktail clinical outcomes study in Covid-19 outpatients. June 6, 2021 (https://www.medrxiv.org/content/10.1101/2021.05.19.21257469v2). preprint.

7. Regeneron’s COVID-19 outpatient trial prospectively demonstrates that REGN-COV2 antibody cocktail significantly reduced virus levels and need for further medical attention. Regeneron, October 28, 2020 (https://investor.regeneron.com/news-releases/news-release-details/regenerons-covid-19-outpatient-trial-prospectively-demonstrates).

8. Baum A, Fulton BO, Wloga E, et al. Antibody cocktail to SARS-CoV-2 spike protein prevents rapid mutational escape seen with individual antibodies. Science 2020; 369: 1014–1018.

9. Copin R, Baum A, Wloga E, et al. The monoclonal antibody combination REGEN-COV protects against SARS-C0V-2 mutational escape in preclinical and human studies. Cell 2021 June 5 (Epub ahead of print).

10. Wang P, Nair MS, Liu L, et al. Antibody resistance of SARS-CoV-2 variants B.1.351 and B.1.1.7. Nature 2021; 593: 130–135.

11. Kanda Y. Investigation of the freely-available easy-to-use software”EZR” (Easy R) for medical statistics. Bone Marrow Transplant. 2013: 48, 452–458. advance online publication 3 December 2012; doi: 10.1038/bmt.2012.244.

12. Hansen J, Baum A, Pascal KE, et al. Studies in humanized mice and convalescent humans yield a SARS-CoV-2 antibody cocktail. Science 2020; 369: 1010–1014.

13. Weinreich DM, Sivapalasingam S, Norton T, et al. REGN-COV2, a neutralizing antibody cocktail, in outpatients with Covid-19. N Engl J Med 2021; 384: 238–251.

14. O’Brien MP, Forleo-Neto E, Musser BJ, et al. Subcutaneous REGEN-COV antibody combination to prevent Covid-19. N Engl J Med 2021 August 4 (Epub ahead of print).

