## Supplementary material for "Impact of Antibody Cocktail Therapy Combined with Casirivimab and Imdevimab on Clinical Outcome for Covid-19 patients in A Real-Life Setting: A Single Institute Analysis": table_figure_supporting information

### Slide 1
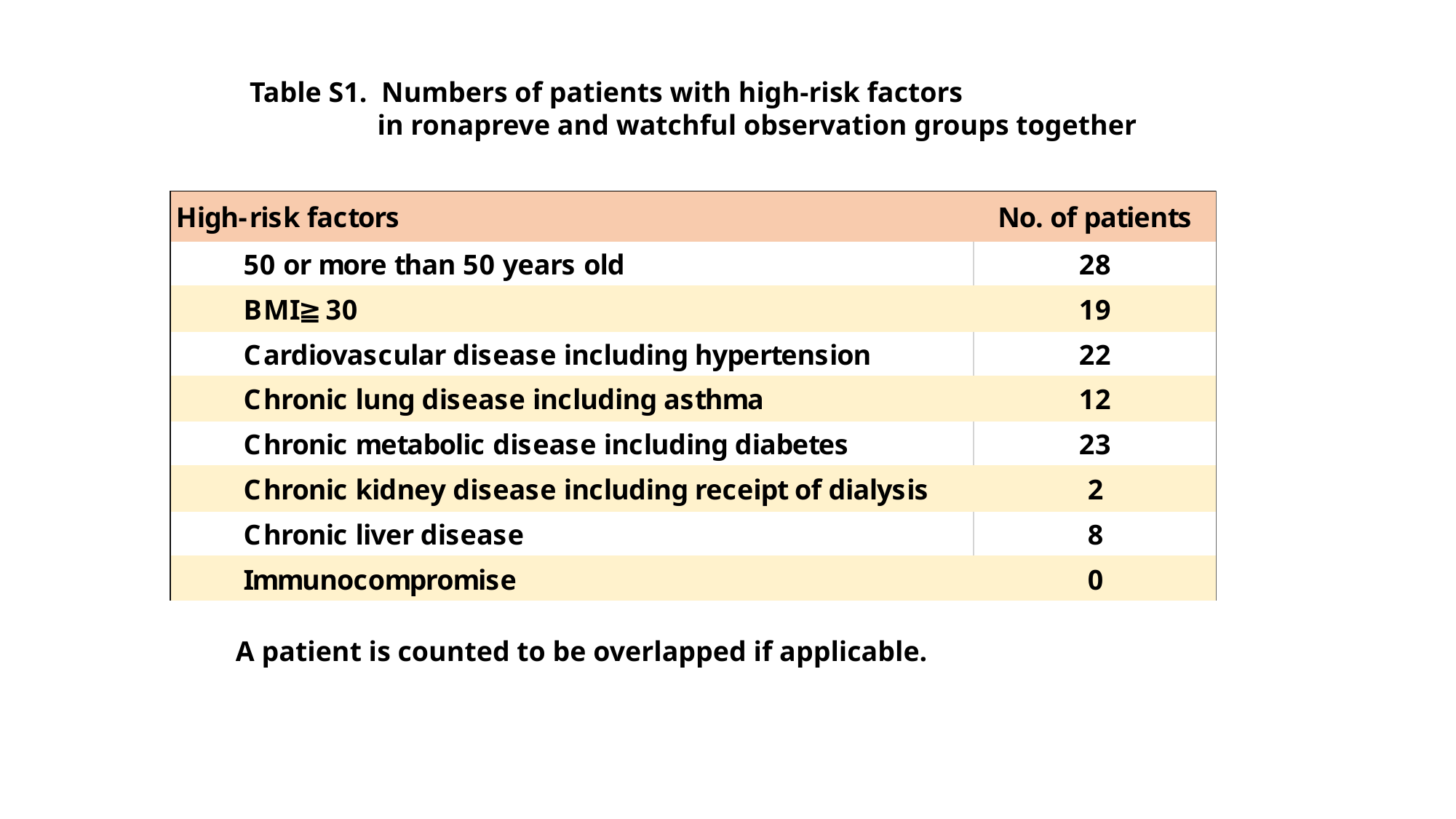

Table S1. Numbers of patients with high-risk factors
 in ronapreve and watchful observation groups together
A patient is counted to be overlapped if applicable.

### Slide 2
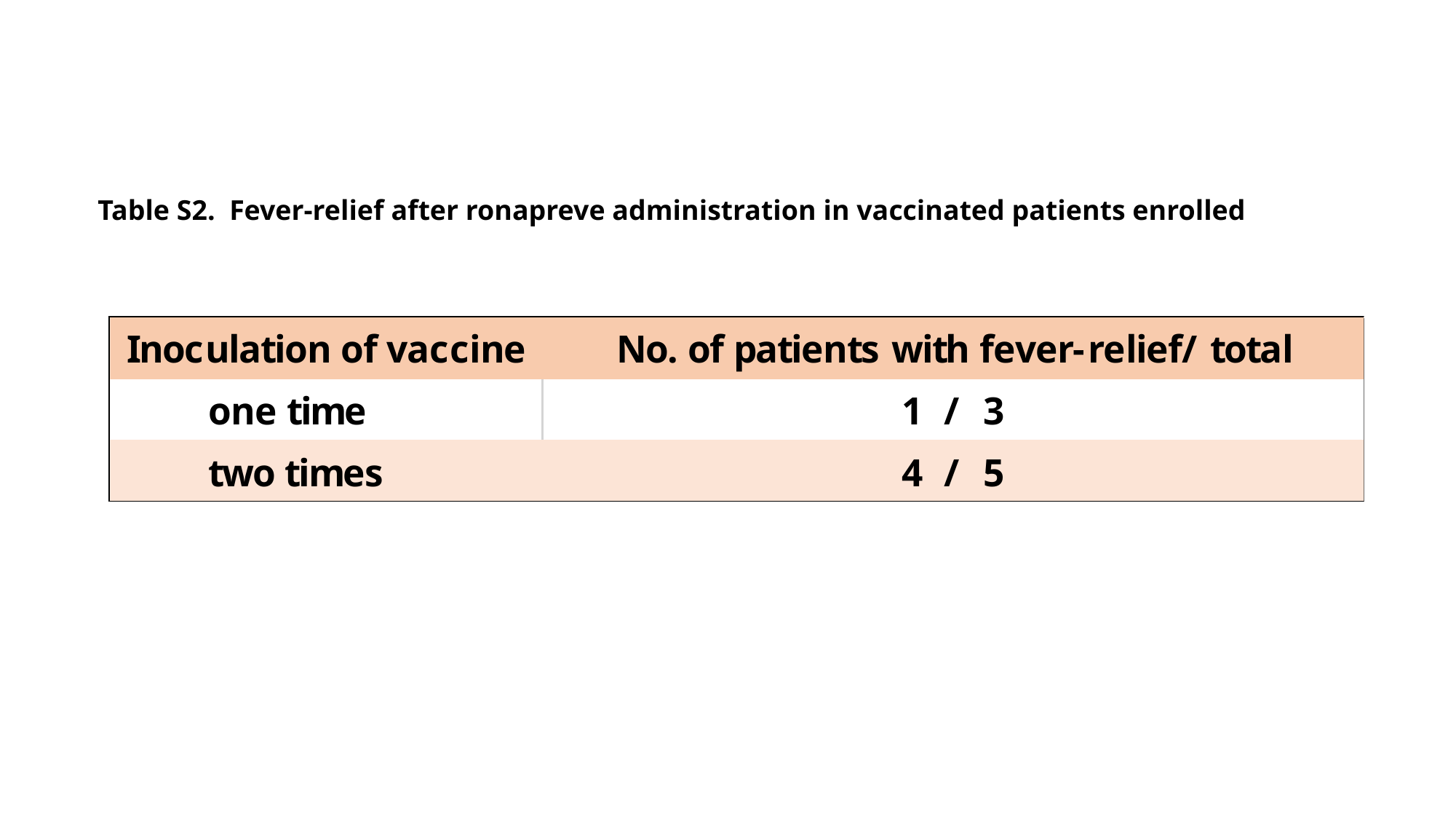

Table S2. Fever-relief after ronapreve administration in vaccinated patients enrolled

### Slide 3
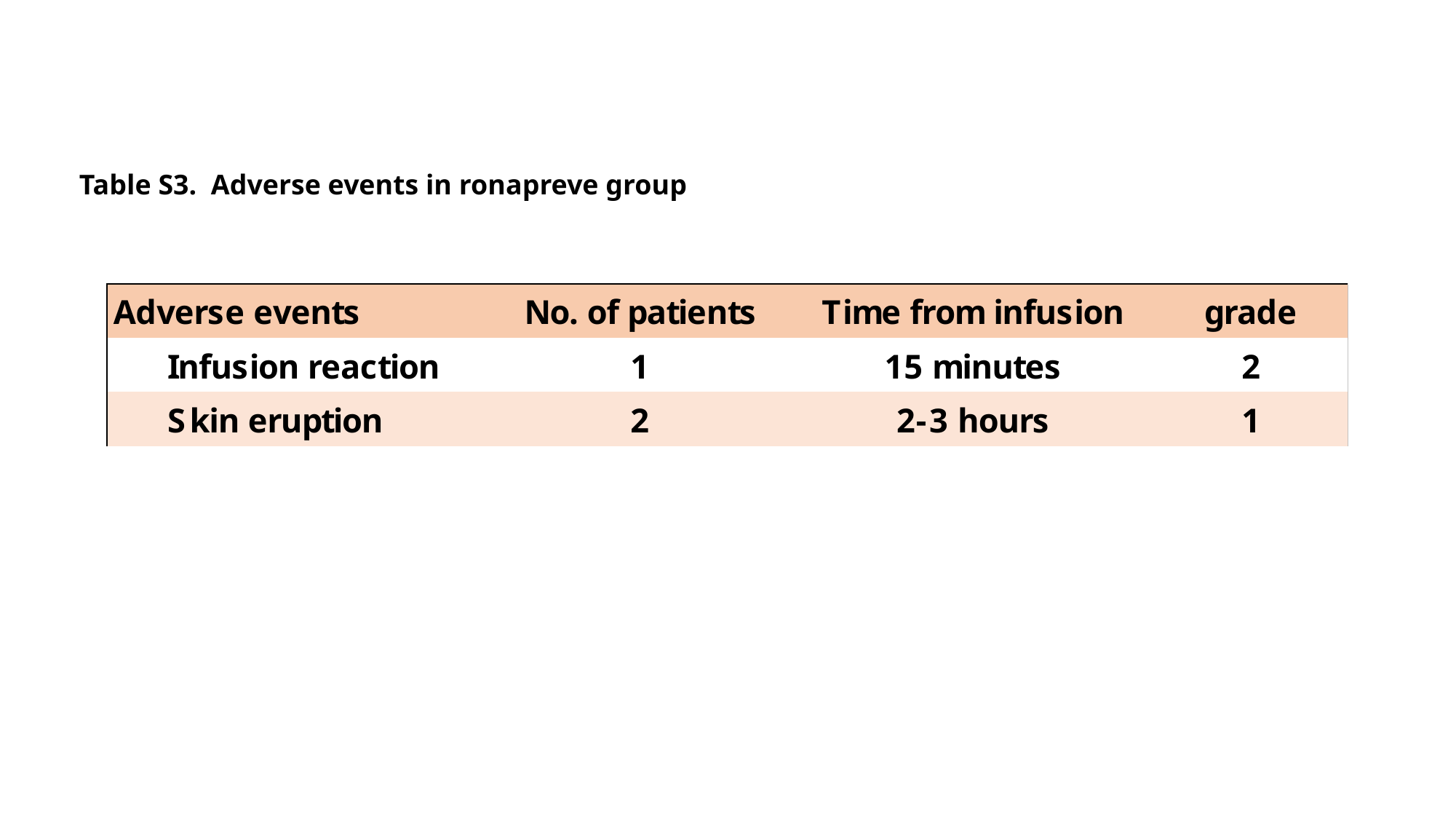

Table S3. Adverse events in ronapreve group

### Slide 4
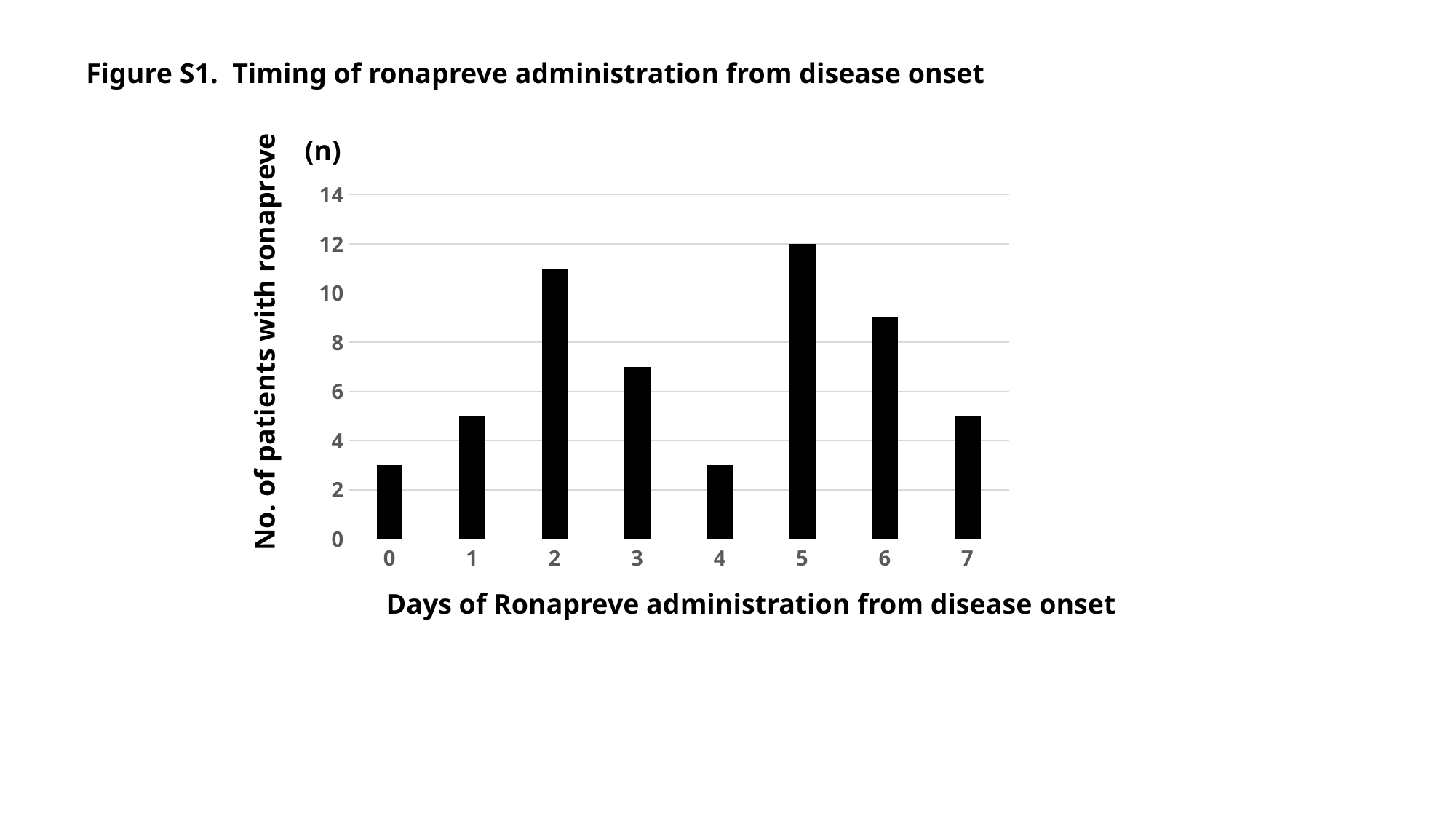

Figure S1. Timing of ronapreve administration from disease onset
(n)
#### Chart
| Category | |
|---|---|
| 0 | 3.0 |
| 1 | 5.0 |
| 2 | 11.0 |
| 3 | 7.0 |
| 4 | 3.0 |
| 5 | 12.0 |
| 6 | 9.0 |
| 7 | 5.0 |No. of patients with ronapreve
Days of Ronapreve administration from disease onset
